# Noninvasive vs Invasive Respiratory Support for Patients with Acute Hypoxemic Respiratory Failure

**DOI:** 10.1101/2023.12.23.23300368

**Authors:** Jarrod M Mosier, Vignesh Subbian, Sarah Pungitore, Devashri Prabhudesai, Patrick Essay, Edward J Bedrick, Jacqueline C. Stocking, Julia M Fisher

**Author notes:** Corresponding Author: Jarrod M. Mosier, MD FCCM Department of Emergency Medicine 1501 N. Campbell Ave., AHSL 4171D PO Box 245057 Tucson, AZ 85724-5057.

## Abstract

**Rationale:** Noninvasive respiratory support modalities are common alternatives to mechanical ventilation for patients with early acute hypoxemic respiratory failure. These modalities include noninvasive positive pressure ventilation, using either continuous or bilevel positive airway pressure, and nasal high flow using a high flow nasal cannula system. However, outcomes data historically compare noninvasive respiratory support to conventional oxygen rather than to mechanical ventilation.

**Objectives:** The goal of this study was to compare the outcomes of in-hospital death and alive discharge in patients with acute hypoxemic respiratory failure when treated initially with noninvasive respiratory support compared to patients treated initially with invasive mechanical ventilation.

**Methods:** We used a validated phenotyping algorithm to classify all patients with eligible International Classification of Diseases codes at a large healthcare network between January 1, 2018 and December 31, 2019 into noninvasive respiratory support and invasive mechanical ventilation cohorts. The primary outcome was time-to-in-hospital death analyzed using an inverse probability of treatment weighted Cox model adjusted for potential confounders, with estimated cumulative incidence curves. Secondary outcomes included time-to-hospital discharge alive. A secondary analysis was conducted to examine potential differences between noninvasive positive pressure ventilation and nasal high flow.

**Results:** During the study period, 3177 patients met inclusion criteria (40% invasive mechanical ventilation, 60% noninvasive respiratory support). Initial noninvasive respiratory support was not associated with a decreased hazard of in-hospital death (HR: 0.65, 95% CI: 0.35 - 1.2), but was associated with an increased hazard of discharge alive (HR: 2.26, 95% CI: 1.92 - 2.67). In-hospital death varied between the nasal high flow (HR 3.27, 95% CI: 1.43 - 7.45) and noninvasive positive pressure ventilation (HR 0.52, 95% CI 0.25 - 1.07), but both were associated with increased likelihood of discharge alive (nasal high flow HR 2.12, 95 CI: 1.25 - 3.57; noninvasive positive pressure ventilation HR 2.29, 95% CI: 1.92 - 2.74),

**Conclusion:** These observational data from a large healthcare network show that noninvasive respiratory support is not associated with reduced hazards of in-hospital death but is associated with hospital discharge alive. There are also potential differences between the noninvasive respiratory support modalities.

## Introduction

Noninvasive respiratory support strategies utilize an external interface (e.g., facemask, helmet, nasal cannula) to deliver either pressure-based support in the form of continuous or bilevel positive airway pressures; or flow-based support in the form of nasal high flow. Noninvasive modalities, particularly pressure-based support, are recommended for acute exacerbations of chronic obstructive pulmonary disease or acute cardiogenic pulmonary edema.(1) Noninvasive respiratory support modalities are also increasingly used for patients with acute *de novo* hypoxemic respiratory failure, despite unclear data on which strategies are superior and safer, as well as the impact on outcomes.(2–5)

Overall, noninvasive strategies likely reduce the need for intubation and consequently lower mortality compared to standard oxygen.(2, 6–8) However, intubation after failed noninvasive respiratory support is associated with prolonged ICU stays and excess mortality.(9–16) The main theory to explain such findings is that nonintubated patients with acute respiratory failure may produce injurious transpulmonary pressures that are inhomogeneously amplified (17) and accelerate lung injury (i.e., patient self-inflicted lung injury). Noninvasive modalities are typically compared to conventional oxygen with the primary outcome most commonly being intubation, either alone or in combination with mortality. The benefits of noninvasive respiratory support (improve respiratory mechanics, reduce work of breathing, and improve gas exchange), however, render noninvasive strategies a more appropriate comparison to mechanical ventilation. Data comparing noninvasive respiratory support to invasive mechanical ventilation after conventional oxygen has proven insufficient, however, are lacking. The goal of this study was to explore that comparison by investigating the outcomes in patients with acute hypoxemic respiratory failure treated with initial noninvasive respiratory support compared to invasive mechanical ventilation.

## Methods

### Study Design, Setting, and Participants

This retrospective cohort study used de-identified structured clinical data from the Banner Health Network clinical data warehouse. Banner Health spans 26 hospitals across six states in the western United States and uses the PowerChart (Cerner Corporation, North Kansas City, MO, USA) electronic health record. Data were extracted for all adult patients (≥18 years) admitted to the hospital between January 1, 2018 and December 31, 2019. Patients were included if they had an admission diagnosis consistent with the pertinent *International Classification of Diseases* (version 10) subcodes with acute hypoxemic respiratory failure (J.96): J96.00, J96.01, J96.02, J96.20, J96.21, J96.22, J96.90, J96.91, and J96.92. Patients were excluded if they had a first treatment location other than emergency department, intensive care unit, stepdown unit, or medical/surgical unit. This work adheres to the STROBE reporting guidelines, guidelines from journal editors,(18) and was approved by the University of Arizona (#1907780973) and Banner Health Institutional Review Boards (#483-20-0018).

### Cohort Assignment

We used a validated phenotyping algorithm to classify eligible cases by the sequence of respiratory support (19–21) into two cohorts: those treated initially with noninvasive respiratory support and those treated with initial invasive mechanical ventilation. All patients in both cohorts were included in the analysis, as there is variation in the determination of failure and physiologic thresholds that prompt intubation.(22) Patients on conventional oxygen only were excluded. Secondary analyses were conducted separating noninvasive respiratory support into noninvasive positive pressure ventilation (either continuous or bilevel positive airway pressure) and nasal high flow. Noninvasive positive pressure ventilation, in any form, in the Banner Health System is provided using a noninvasive ventilator, and nasal high flow is delivered by either the Vapotherm (Vapotherm, Exeter, New Hampshire) system or the OptiFlow (Fisher & Paykel, Auckland, New Zealand) with or without the AirVo2 system.

A subset of patients received noninvasive positive pressure ventilation, nasal high flow, and invasive mechanical ventilation. These patients were manually assigned to the noninvasive respiratory support or invasive mechanical ventilation cohort based on treatment start times for analyses comparing noninvasive support to mechanical ventilation but were excluded from analyses separating noninvasive support into noninvasive positive pressure ventilation and nasal high flow.

We estimated the propensity for invasive mechanical ventilation or noninvasive respiratory support (noninvasive positive pressure ventilation or nasal high flow separately in secondary analyses) by using generalized boosted models and used inverse probability of treatment weighting in the models to account for non-random treatment assignment,(23) mirroring our previous comparisons in patients with COVID-19 associated respiratory failure.(20) The variables for propensity score estimation included age, body mass index, sex, ethnicity (non-Hispanic, Hispanic), race (white, other), respiratory rate and SpO_2_/FiO_2_ ratio immediately prior to first treatment, comorbidities (diabetes, chronic kidney disease, heart failure, hypertension, chronic obstructive pulmonary disease, neoplasm/immunosuppression, chronic liver disease, obesity), diagnoses of influenza or sepsis, vasopressor infusion before first treatment, first treatment location (emergency department, intensive care unit, stepdown, med/surg), hospital, time period of hospital admission (time period 1 [January 1 - June 30, 2018], time period 2 [July 1 – December 31, 2018], time period 3 [January 1 - June 30, 2019], and time period 4 [July 1 - December 31, 2019]), and hours from hospital admission to first treatment, transformed via the Box-Cox method with negatives.(24) Hospitals with <30 observations were grouped together for ease of modeling and to preserve de-identification. These variables were additionally included in later modeling to further improve balance between treatment groups.

### Outcomes and Data Analysis

The primary outcome was time-to-in-hospital death, defined as time from initiation of respiratory support to death with hospital discharge considered a competing event. It was modeled using a cause-specific Cox model with the first treatment (noninvasive respiratory support versus invasive mechanical ventilation) as the key predictor. A secondary outcome of time-to-hospital discharge alive was also evaluated using the same method. Each outcome was also evaluated with secondary analyses separating noninvasive respiratory support into noninvasive positive pressure ventilation and nasal high flow and a sensitivity analysis that removed patients with evidence of all three treatments. We assessed the proportional hazard assumption in the Cox models by including an interaction of time with first treatment and reported the model with the interaction if it was statistically significant at α = 0.05. Then, because of the limitations of hazard ratios in the presence of competing events, we estimated cumulative incidence curves associated with each treatment using the Cox model estimates.(25) We explored cumulative incidence curves associated with the comorbidities heart failure and chronic obstructive pulmonary disease and diagnoses of influenza or sepsis both individually and in combination. We set the remaining covariate values to their sample median (continuous covariates) or most frequent value (categorical covariates).(26, 27) The unweighted outcomes of mortality, intubation rate, days to intubation, and duration of mechanical ventilation were assessed using Fisher’s Exact and Kruskal-Wallis rank sum tests where appropriate.

Electronic health record data requires accounting for varying levels of missingness among variables.(28–30) Missing data were handled by using multiple imputation by chained equations.(31, 32) For each analysis, we created 50 imputed data sets using all variables in the propensity score, the Nelson-Aalen estimate of the cumulative hazard rate function of available time-to-event data, the time-to-event itself, and event information (i.e., in-hospital death, hospital discharge alive). Body mass index, SpO_2_/FiO_2_, and respiratory rate were imputed via predictive mean matching. Sex, ethnicity, race, and comorbidities were imputed with logistic regression. All variables used in the propensity score estimation and the outcome variables were used to model any variables with missing data with the exception that raw time-to-event was not used to predict other variables in the multiple imputation by chained equations algorithm. Instead, temporal information was used in the prediction of missing values via the Nelson-Aalen estimate. We estimated propensity scores for each imputed data set separately. For the Cox models, the propensity scores from a specific imputed data set were used for inverse probability of treatment weighting for that data set,(31, 32) and results were combined using Rubin’s Rules. All data preprocessing and statistical analyses were done using R version 4.1.0(33) and included the following packages: twang(34), survival (35, 36), survminer,(37) mice (31), xtable,(38) and tidyverse.(39) Further detailed descriptions of data preprocessing can be found our previous work.(20)

## Results

There were 3177 patients who met the inclusion criteria. Of these, 1266 (40%) were intubated initially while 1911 (60%) were initially treated with noninvasive respiratory support, **Figure 1**, **Table 1**. There are important differences between cohorts. Patients intubated initially were more commonly male (55% vs. 49%) and disproportionately at large hospitals (57% vs. 44%) compared to patients initially treated with noninvasive respiratory support. They were also a higher acuity based on median APACHE score (68 vs. 49), although only patients admitted to an intensive care unit were given an APACHE score in the electronic health record. Those intubated initially were less likely to have comorbid heart failure (36% vs. 47%) or chronic obstructive pulmonary disease (68% vs. 85%), more likely to be septic (36% vs. 19%), and had more severe hypoxemia based on SpO_2_/FiO_2_ on treatment assignment (medians 130 vs. 261, difference of means 90.52, 95% CI: 84.04 - 97.01) despite clinically similar worst PaO_2_/FiO_2_ in the first 24 hours (medians 124 vs. 142, difference of means 4.59, 95% CI: -4.39 - 13.58). Of the 1850 (97%) patients where the sequence of noninvasive respiratory support could be reliably classified, most patients (96%) were treated with noninvasive positive pressure ventilation (supplementary **Table E1**). All-cause in-hospital mortality was 13%, higher for patients intubated first than for those treated with noninvasive respiratory support first (18% vs 9%), **Table 2**. Mortality was significantly higher for noninvasive respiratory support patients who required intubation compared to those who didn’t (22% vs. 6%).

**Figure 1:**
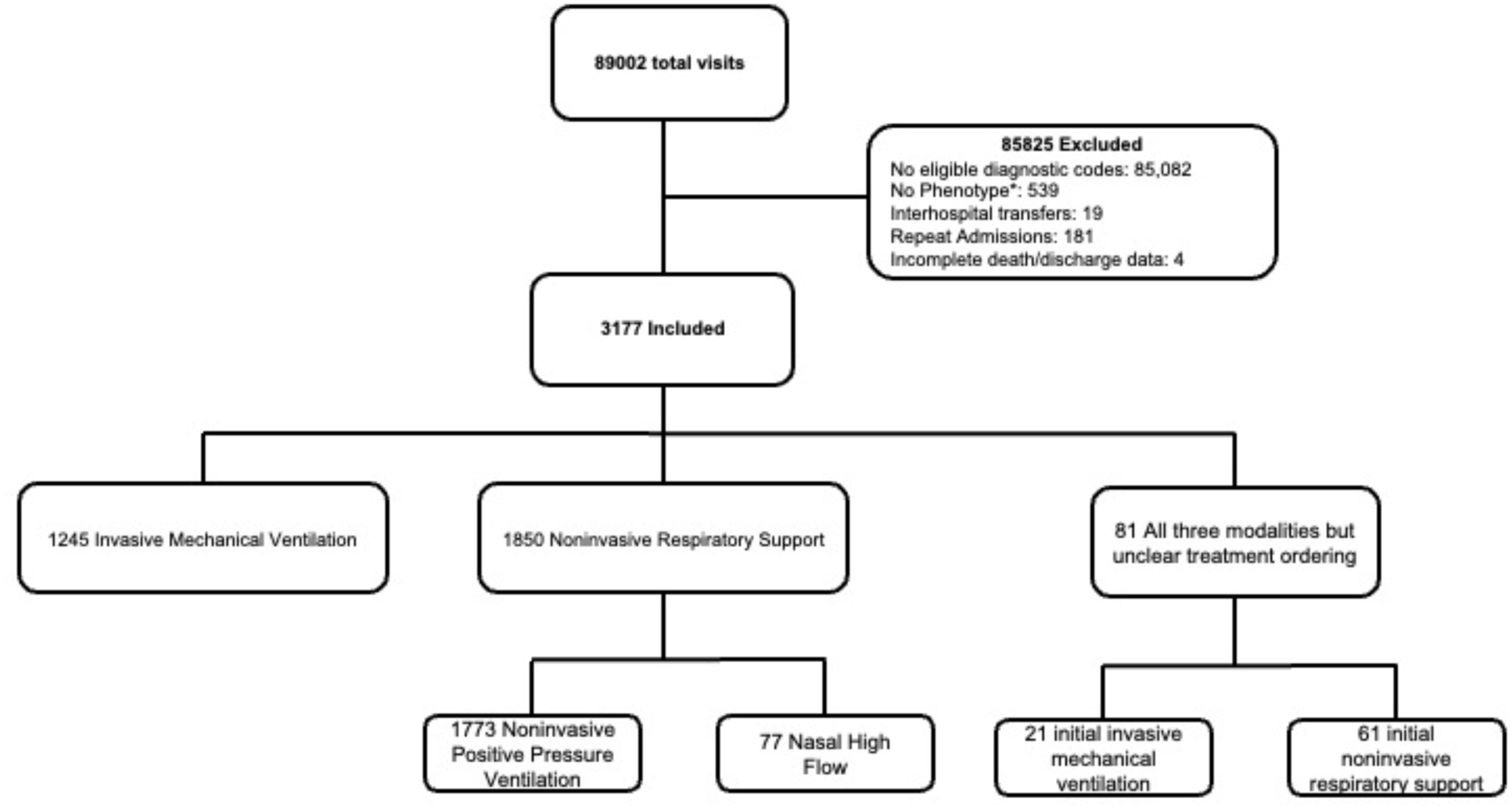
STROBE Statement. Figure Caption: STROBE diagram of included subjects. There were 89,002 total visits during the study period. Of those, most (85,825) failed to meet exclusion criteria. *The subjects that were excluded because they were not classified by the algorithm but had an eligible diagnostic code on admission likely represent those only requiring conventional oxygen. Repeat admissions and interhospital transfers were excluded due to confounding with the outcome.

**Table 1:** Demographics.

| Measure | Invasive Mechanical Ventilation | Noninvasive Respiratory Support | Total |
| --- | --- | --- | --- |
| N (%) | 1266 (40%) | 1911 (60%) | 3177 |
| Female Sex | 574 (45%) | 970 (51%) | 1544 (49%) |
| Age, median (IQR) | 61 (48 - 72) | 68 (58 - 77) | 66 (54 - 75) |
| BMI, median (IQR) | 28 (23 - 35) | 29 (23 - 37) | 28 (23 - 36) |
| Ethnicity, n(%)** |  |  |  |
| Not Hispanic or Latino | 1031 (82%) | 1645 (86%) | 2676 (85%) |
| Hispanic or Latino | 225 (18%) | 261 (14%) | 486 (15%) |
| Race, n (%)** |  |  |  |
| White | 1078 (82%) | 1690 (89%) | 2768 (88%) |
| Black or African American | 76 (6%) | 105 (6%) | 181 (6%) |
| Asian/Native Hawaiian/Pacific Islander | 16 (1%) | 18 (1%) | 34 (1%) |
| American Indian or Alaska Native | 62 (5%) | 39 (2%) | 101 (3%) |
| Other | 25 (2%) | 53 (3%) | 78 (2%) |
| Hospital Size<sup>a</sup>, n(%) |  |  |  |
| small | 39 (4%) | 158 (9%) | 197 (7%) |
| medium | 428 (39%) | 815 (47%) | 1243 (44%) |
| large | 631 (57%) | 779 (44%) | 1410 (49%) |
| APACHE IVa median (IQR) | 68 (50 - 87) | 49 (38 - 63) | 57 (43 - 76) |
| Vital Signs on Treatment Assignment median (IQR) |  |  |  |
| Heart rate | 93 (79 - 112) | 88 (75 - 104) | 90 (76 - 107) |
| Systolic blood pressure | 120 (103 - 140) | 132 (116 - 149) | 127 (111 - 146) |
| Diastolic blood pressure | 68 (57 - 82) | 73 (64 - 83) | 71 (61 - 83) |
| SpO2 | 98 (96 - 100) | 96 (93 - 98) | 97 (94 - 99) |
| FiO2<sup>b</sup> | 75 (50 - 100) | 36 (30 - 50) | 45 (33 - 80) |
| SpO2:FiO2 | 130 (99-200) | 261 (186 - 323) | 200 (116 - 297) |
| Temperature (°C) | 37 (36.6 - 37) | 36.8 (36.5 - 37) | 36.9 (36.5 - 37) |
| Respiratory Rate | 19 (16 - 23) | 20 (18 - 25) | 20 (18 - 24) |
| Comorbidities<sup>c</sup> n(%) |  |  |  |
| Diabetes | 468 (40%) | 756 (40%) | 1224 (40%) |
| Chronic Kidney Disease | 261 (22%) | 527 (28%) | 788 (26%) |
| Heart Failure | 418 (36%) | 903 (47%) | 1321 (43%) |
| Hypertension | 839 (72%) | 1427 (75%) | 2266 (74%) |
| Chronic Liver Disease | 242 (21%) | 157 (8%) | 399 (13%) |
| Neoplasm or Immunosuppression | 175 (15%) | 293 (15%) | 468 (15%) |
| COPD | 799 (68%) | 1613 (85%) | 2412 (78%) |
| Obesity | 186 (16%) | 350 (18%) | 536 (17%) |
| Acute Influenza Diagnosis | 2 (0%) | 4 (0%) | 6 (0%) |
| Acute sepsis diagnosis | 419 (36%) | 354 (19%) | 773 (25%) |
| Labs on Admission median (IQR) |  |  |  |
| PaO2 (mmHg) (Worst Value) | 79 (65 - 107) | 75 (63 - 102) | 77 (63 - 104) |
| PaO2:FiO2 (Worst Value) | 124 (79 - 220) | 142 (84 - 218) | 133 (81 - 218) |
| White Blood Cell Count (K/uL) | 8 (2.45 - 14) | 7.7 (2 - 13) | 7.9 (2 - 13.5) |
| Lactate (mmol/L) | 1.9 (1.2 - 3.7) | 1.5 (1 - 2.3) | 1.7 (1.1 - 2.8) |
| pH | 7.31 (7.19 - 7.39) | 7.33 (7.26 - 7.41) | 7.33 (7.24 - 7.4) |
| PaCO2 (mmHg) | 46 (38 - 64) | 53 (40 - 70) | 50 (38 - 68) |
| HCO3 (mmol/L) | 24 (20 - 28) | 27 (23 - 32) | 26 (22 - 30) |
| BNP (pg/mL) | 1490 (359 - 5781) | 1316 (327 - 5381) | 1369 (334 - 5476) |
| Creatinine (mg/dL) | 1.0 (0.76 - 1.58) | 0.95 (0.73 - 1.36) | 0.97 (0.74 - 1.43) |
| Time from hospital admission to treatment (h) | 0.6 (0 - 7) | 27 (0 - 89) | 6 (0 - 61) |
| Treatment Assignment Location n (%)** |  |  |  |
| Emergency Department | 666 (53%) | 523 (27%) | 1189 (37%) |
| ICU | 505 (40%) | 644 (34%) | 1149 (36%) |
| Non-ICU ward | 16 (1%) | 272 (14%) | 288 (9%) |
| Stepdown | 79 (6%) | 472 (25%) | 551 (17%) |

\*\*Data are presented as percent of available.
<sup>b</sup>FiO2 determined by documented FiO2, if documented, or by $FiO_2 = 100(0.21 + \text{oxygen flow [L/min}^{-1}] \times 0.03$ if a flow rate was documented.

**Table 2:** Unmatched outcomes.

| Outcome | Invasive Mechanical Ventilation | Noninvasive Respiratory Support | Total | P-Value |
| --- | --- | --- | --- | --- |
| All-Cause Hospital Mortality<br>Total<br>Failure<br>Success | 239 (19% of 1266) | 201 (11% of 1911)<br>105 (27% of 384)<br>96 (6% of 1527) | 440 (14% of 3177) | p < 0.001<br>p < 0.001* |
| Failure (intubation rate) |  | 384 (20% of 1911) |  |  |
| Days to Intubation, median (IQR)<br>From starting noninvasive respiratory support<br>From hospital admission | --<br>0.03 (-0.03 - 0.3) | 0.15 (0.07 - 0.35)<br>0.51 (0.05 - 3.64) | --<br>0.05 (-0.02 - 0.87) | --<br>p < 0.001 |
| Duration of Mechanical Ventilation, days, median (IQR) | 1.61 (0.67 - 4.47) | 2.17 (0.88 - 5.49) | 1.77 (0.71 - 4.67) | p = 0.002 |
| Hospital Length-of-Stay<br>Total<br>Failure<br>Success | 6.89 (3.59 - 13.02) | 5.31 (3.1 - 9)<br>8.21 (4.48 - 15.84)<br>4.97 (2.96 - 8) | 5.85 (3.25 - 10.14) | p < 0.001<br>p < 0.001* |
| Estimates are n (% of column n with available data) for categorical characteristics and median (interquartile range) for continuous characteristics. Inferences for categorical variables are the result of Fisher Exact Tests, with p-values computed via Monte Carlo simulation when necessary for computational efficiency. Inferences for continuous variables come from Kruskal-Wallis rank sum test of group differences. *These p-values correspond to tests of outcome distribution differences by failure/success among NIRS patients. |  |  |  |  |

Hazard ratios are shown in **Table 3**. Initial noninvasive respiratory support was not associated with a decreased hazard of in-hospital death (HR: 0.65, 95% CI: 0.35 - 1.2); with no significant interaction of treatment and time and a reasonable protection from unmeasured confounders with an E-value of 2.04. Initial noninvasive respiratory support was, however, associated with an increased hazard of discharge alive (HR: 2.26, 95% CI: 1.92 - 2.67) that decreased over time (interaction between time and noninvasive respiratory support HR: 0.97, 95% CI: 0.95 - 0.98) and an even stronger E-value (2.90). Sensitivity analyses excluding the patients without clear treatment sequence shows noninvasive respiratory support was associated with a reduced hazard of in-hospital death (HR: 0.53, 95% CI: 0.28 - 0.99, E-value 2.49) and an increased hazard of hospital discharge alive (HR: 2.34, 95% CI: 1.97 - 2.77, E-value 2.98) that again decreased over time (interaction between time and noninvasive respiratory support HR: 0.97, 95% CI: 0.95 - 0.98). Representative cumulative incidence curves show a consistent trend of slightly reduced probability of in-hospital death and increased probability of discharge alive for noninvasive respiratory support across a range of comorbidities (chronic obstructive pulmonary disease, congestive heart failure) and diagnoses (influenza, sepsis), **Supplementary Figures E1, E2**.

**Table 3:**
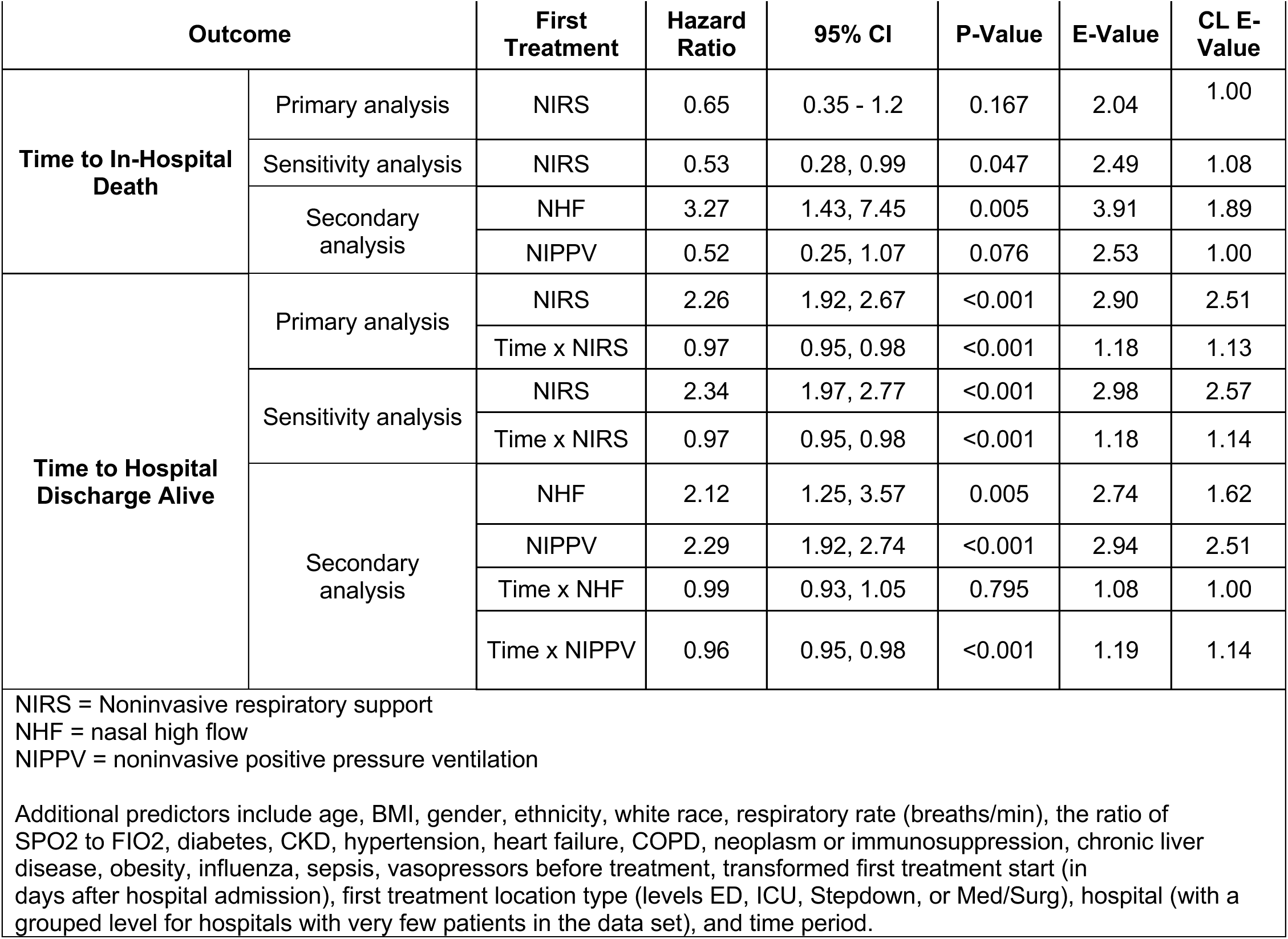
Cox Model Results.

**Figure 2:**
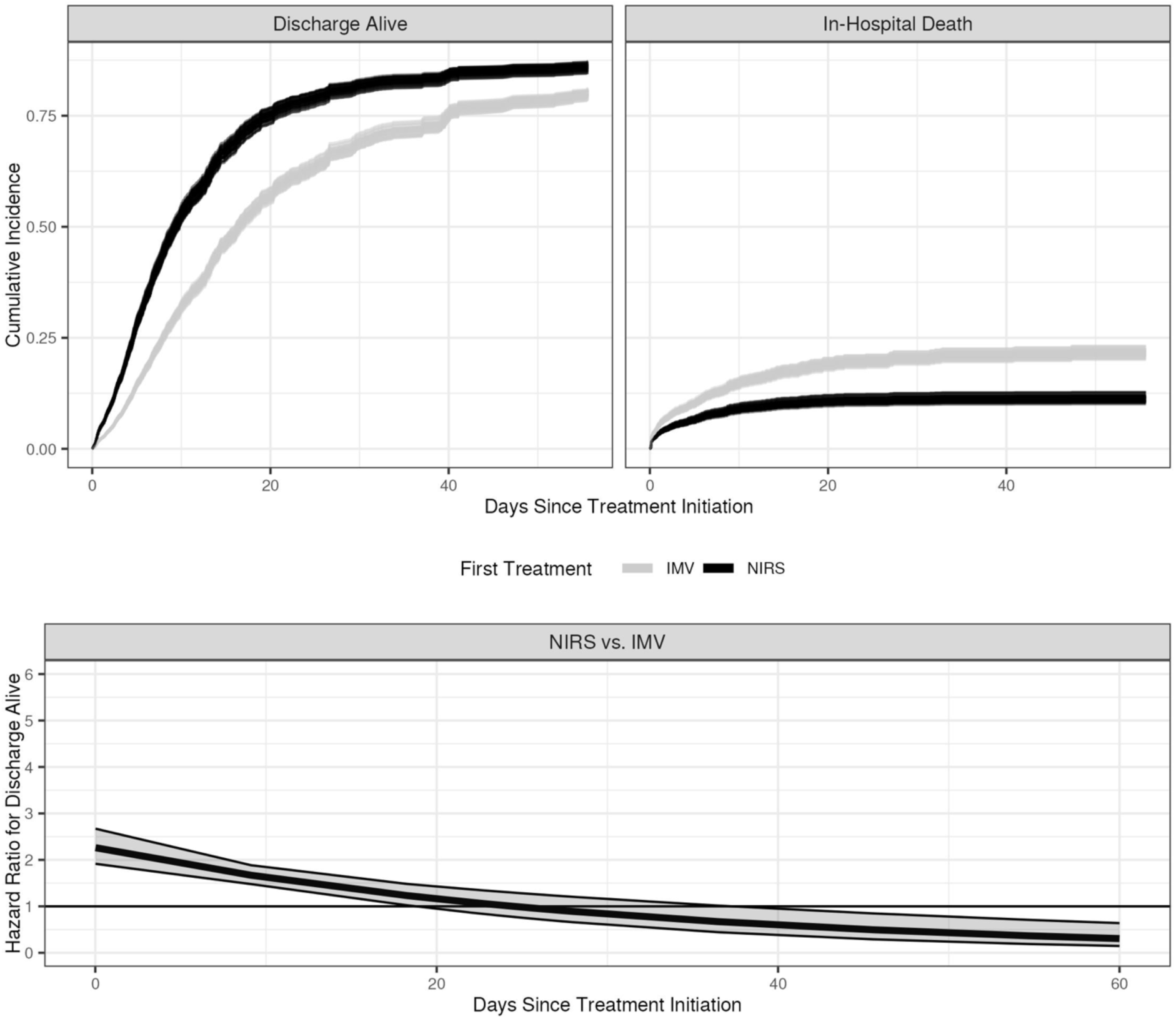
Top: Model-estimated cumulative incidence curves for noninvasive respiratory support (NIRS) vs invasive mechanical ventilation (IMV) showing the probabilities for hospital discharge alive (left) and in-hospital death (right). Bottom: Estimated time-varying hospital discharge alive hazard ratios for NIRS versus IMV with pointwise 95% confidence intervals. The following values were used for covariates: male, not Hispanic or Latino, white, one of the large hospitals (hospital A), hospital admission to the emergency department between January 1, 2018 and June 30, 2018, no vasopressor infusion before treatment, no diabetes, no chronic kidney disease, no heart failure, yes hypertension, no chronic obstructive pulmonary disease, no neoplasm/immunosuppression, no chronic liver disease, no obesity, no influenza, yes sepsis, and continuous covariates set at their median values (age = 66 years, SpO_2_/FiO_2_ = 200, respiratory rate = 20 breaths/min, BMI = 28.44, transformed hours from hospital admission to first treatment = 1.77). Each imputed data set generates a pair of curves (one for IMV, one for NIRS). The probability of discharge alive is greater for NIRS than for IMV, and the probability of in-hospital death is lower. The hazard ratio of discharge alive for NIRS vs. IMV starts out statistically significantly positive shortly before 20 days after treatment initiation and eventually switches direction around 40 days.

**Figure 3:**
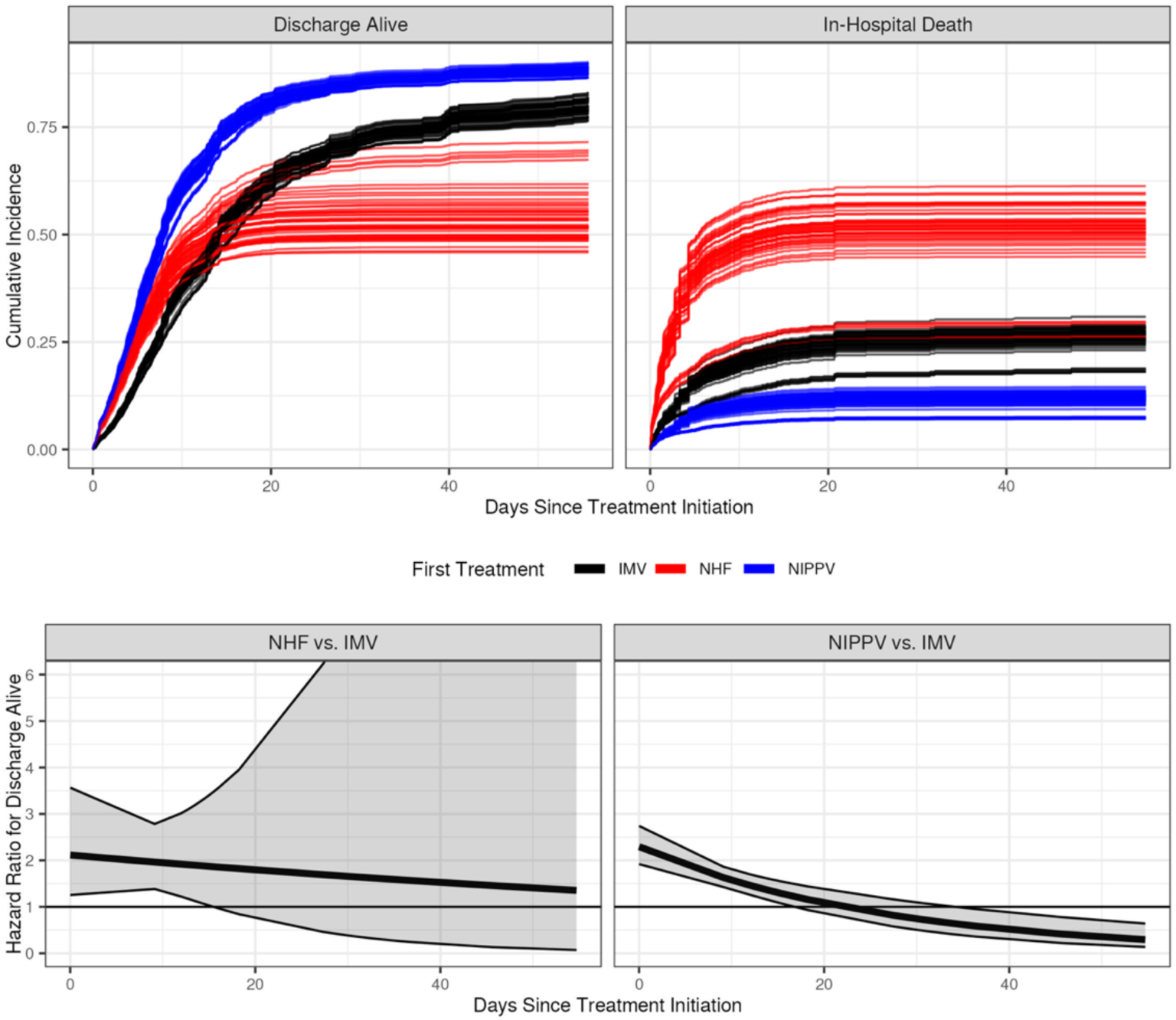
Top: Model-estimated cumulative incidence curves for noninvasive positive pressure ventilation (NIPPV), nasal high flow (NHF), and invasive mechanical ventilation (IMV) showing the probabilities for hospital discharge alive (left) and in-hospital death (right). Bottom: Estimated time-varying hospital discharge alive hazard ratios for NHF versus IMV (left) and NIPPV versus IMV (right) with pointwise 95% confidence intervals. The following values were used for covariates: male, not Hispanic or Latino, white, one of the large hospitals (hospital A), hospital admission to the emergency department between January 1, 2018 and June 30, 2018, no vasopressor infusion before treatment, no diabetes, no chronic kidney disease, no heart failure, yes hypertension, no chronic obstructive pulmonary disease, no neoplasm/immunosuppression, no chronic liver disease, no obesity, no influenza, yes sepsis, and continuous covariates set at their median values (age = 66 years, SpO_2_/FiO_2_ = 200, respiratory rate = 20 breaths/min, BMI = 28.44, transformed hours from hospital admission to first treatment = 1.70). Each imputed data set generates a triple of curves (one for IMV, NHF, and NIPPV). Patients initially treated with either non-invasive modality had a higher probability of hospital discharge alive until roughly 15 days after treatment initiation. After that, the probability of discharge alive remained higher for NIPPV compared to IMV but reversed direction for NHF and IMV. Nasal high flow had a slightly higher probability of in-hospital death than IMV, and NIPPV had a slightly lower probability of in-hospital death than IMV. The hospital discharge alive hazard ratio of NHF to IMV was positive but decreasing until around day 15, at which point there was no further clear difference between those two treatments. The same pattern held for the hospital discharge alive hazard ratio of NIPPV to IMV except that it eventually reversed direction, resulting in the hazard of hospital discharge alive being greater for IMV than NIPPV starting at roughly 35 days after treatment initiation.

In the secondary analyses separating noninvasive respiratory support modalities, the hazard for in-hospital death ranged from increased with nasal high flow (HR 3.27, 95% CI: 1.43 - 7.45, E-value 3.91) to non-significantly decreased with noninvasive positive pressure ventilation (HR 0.52, 95% CI 0.25 - 1.07, E-value 2.53), neither with an interaction with time. Both modalities were associated with increased hazard of discharge alive (nasal high flow HR 2.12, 95 CI: 1.25 - 3.57, E-value 2.74; noninvasive positive pressure ventilation HR 2.29, 95% CI: 1.92 - 2.74, E-value 2.94), both with strong E-values, but only noninvasive positive pressure ventilation showed an interaction with time (interaction between time and nasal high flow HR 0.99, 95% CI: 0.93 - 1.05; interaction between time and noninvasive positive pressure ventilation HR 0.96, 95% CI: 0.95 - 0.98). Representative cumulative incidence curves are shown in the **online supplement Figures E3 & E4**.

## Discussion

Noninvasive respiratory support strategies are increasingly used as alternative initial strategies to early intubation and mechanical ventilation for patients with acute hypoxemic respiratory failure. Thus, the goal of this study was to compare the outcomes between those two approaches. Our results show that initial noninvasive respiratory support modalities in patients with acute hypoxemic respiratory failure were most likely not associated with a reduced hazard of in-hospital death (no association in the primary analyses, weakly significant association in the sensitivity analysis), yet were associated with an increased probability of hospital discharge alive when compared to initial invasive mechanical ventilation. The existing literature comparing noninvasive strategies to conventional oxygen suggests that noninvasive respiratory support is probably associated with reduced mortality, reduced intubation, and shorter hospitals stays, but to varying degrees among the different noninvasive modalities.(2, 4, 5, 40, 41) Our results expand upon this knowledge by comparing outcomes between noninvasive strategies and invasive mechanical ventilation in non-COVID-19 acute hypoxemic respiratory failure. Despite patients who were intubated first generally being more severely ill and more likely to be septic, there was no increase in hazard of our primary outcome (in-hospital death). However, noninvasive respiratory support was associated with an increased likelihood of hospital discharge alive. Further study may show that failure of a noninvasive respiratory support modality may be associated with increased mortality beyond the progression of disease, thus having an outsized influence on the overall association with mortality. This hypothesis is supported by the observed 3.5-fold increase in unweighted mortality for patients that failed a noninvasive strategy compared to those that did not.

These data suggest that noninvasive respiratory support modalities can be effective alternatives to mechanical ventilation for the initial treatment of some patients with acute hypoxemic respiratory failure. Successful noninvasive support increases the likelihood of earlier hospital discharge, but an unsuccessful trial may carry outsized consequences for mortality. These results add to findings from the Lung Safe study, which showed that noninvasive positive pressure ventilation was used in 15% of patients with acute respiratory distress syndrome, but both failure and mortality increased as the severity of disease worsened.(15) While spontaneous breathing can have some advantages in acute hypoxemic respiratory failure patients, patient self-inflicted lung injury is the likely reason for the worse outcomes in patients that fail a noninvasive strategy.(42, 43)

Our secondary analyses also suggested differences in outcomes between noninvasive positive pressure ventilation and nasal high flow. There are four possible explanations for these findings. The first possibility is that noninvasive positive pressure may be the better noninvasive strategy. The second possibility is that the patients treated with nasal high flow may not have been similar to the patients treated with noninvasive positive pressure ventilation, and that our efforts to account for treatment confounding were not fully successful. Patients treated with nasal high flow in our dataset had a higher median APACHE score on admission (56 vs. 48, difference of means 11.77, 95% CI: 6.36, 17.18) a lower median SpO_2_/FiO_2_ on treatment assignment (136 vs 274, difference of means -105, 95% CI: -126 - -85) and lower median worst PaO_2_/FiO_2_ (76 vs 150, difference of means -76, 95% CI: -106 - -45), more commonly had neoplasm or immunosuppression (26% vs 15%) and were more commonly septic (34% vs 17%). The third possible explanation is that patients may not have been treated similarly. Patients initiated on noninvasive positive pressure in a non-intensive care unit were more commonly transferred to the intensive care unit by 12 hours than patients started on nasal high flow (32% vs 12%). Lastly, there may have been imbalanced, imprecise, or incorrect delivery of one modality compared to the other. The median flow rate for nasal high flow was 40lpm (95% CI: 35 - 50lpm). While gas exchange can improve at lower flow rates, higher flow rates are required for the work of breathing benefits related to changes in resting lung volume and strain.(44) Monitoring likely differed between intensive care (e.g., work of breathing changes, signs of fatigue) and non-intensive care units (e.g., oxygen saturation). Additionally, managing failure could have differed as failing nasal high flow could have resulted in more crossover to noninvasive positive pressure ventilation than intubation compared to failing noninvasive positive pressure ventilation.

There are important limitations to our data. Our data were limited to pre-COVID-19 data. As such, the use of noninvasive respiratory support modalities has likely evolved as is evident by the relatively low number of patients treated with nasal high flow across the entire health network. Second, we used admission diagnostic codes to select patients treated at the time for acute hypoxemic respiratory failure without the selection bias of using discharge diagnostic codes. As such, these results are contingent upon accurate coding of admission diagnoses and important patient groups may have been excluded. Since these data are non-randomized observational data, non-protocolized clinical care may have contributed unmeasured confounding differences in the selection for and management of each modality. We attempted to control for confounding by inverse probability for treatment assignment weighting and further adjusting for potential confounders in the Cox models. Furthermore, our E-values are relatively strong, so any unmeasured confounding would have needed to have a substantial effect to alter the results. Another limitation is that results are based on the first assigned therapy, and symptom onset time is not available in our dataset. Thus, crossover (and imbalanced crossover), and symptom duration could confound the findings. Lastly, goals of care and end-of-life issues are not extractable from structured electronic health record data. There is an important difference between a patient who is a do-not-intubate on admission treated with rescue noninvasive respiratory support and a similar patient who worsened during noninvasive respiratory support and chose to become a do-not-intubate and elect comfort focused treatment. However, both patients would have been included in our dataset and could contribute some confounding in the results.

Despite these limitations, our results provide an overview of outcomes between respiratory support modalities that were pragmatically applied for patients with acute hypoxemic respiratory failure across a large healthcare network. These results highlight important knowledge gaps needing further study, including: 1. the risks of failing noninvasive respiratory support, mechanisms of those risks, thresholds of, monitoring for, and management of failure, 2. reproducible phenotypes likely to do well or not do well with noninvasive respiratory support modality, 3. optimal noninvasive support modality by phenotype, 4. optimal noninvasive respiratory support delivery by modality and, 5. optimal hospital location and minimal monitoring capabilities for patients with acute hypoxemic respiratory failure requiring noninvasive respiratory support.

Our data across a large and diverse healthcare network show that initial treatment with noninvasive respiratory support is not associated with a reduced hazard of in-hospital death compared to invasive mechanical ventilation for patients admitted with acute hypoxemic respiratory failure. However, noninvasive respiratory support is associated with a higher likelihood of earlier hospital discharge. Lastly, our data suggest potential differences between noninvasive respiratory support modalities that require further exploration.

## Data Availability

Data are unavailable

## Author Contributions

JMM, VS, JMF, and PE conceived the study idea. PE, VS, and JMM developed the phenotyping algorithm. PE, DP, JMF, JMM, and SP preprocessed the data. PE and VS applied the phenotyping algorithm. JMF, DP, EJB, and SP performed the statistical analysis. JMM and JMF drafted the initial manuscript, and all authors participated in manuscript revisions.

## Acknowledgements

The authors would like to thank Don Saner and Mario Arteaga from the Banner Health Network Clinical Data Warehouse for their support during this project.

---

**Table E1:**
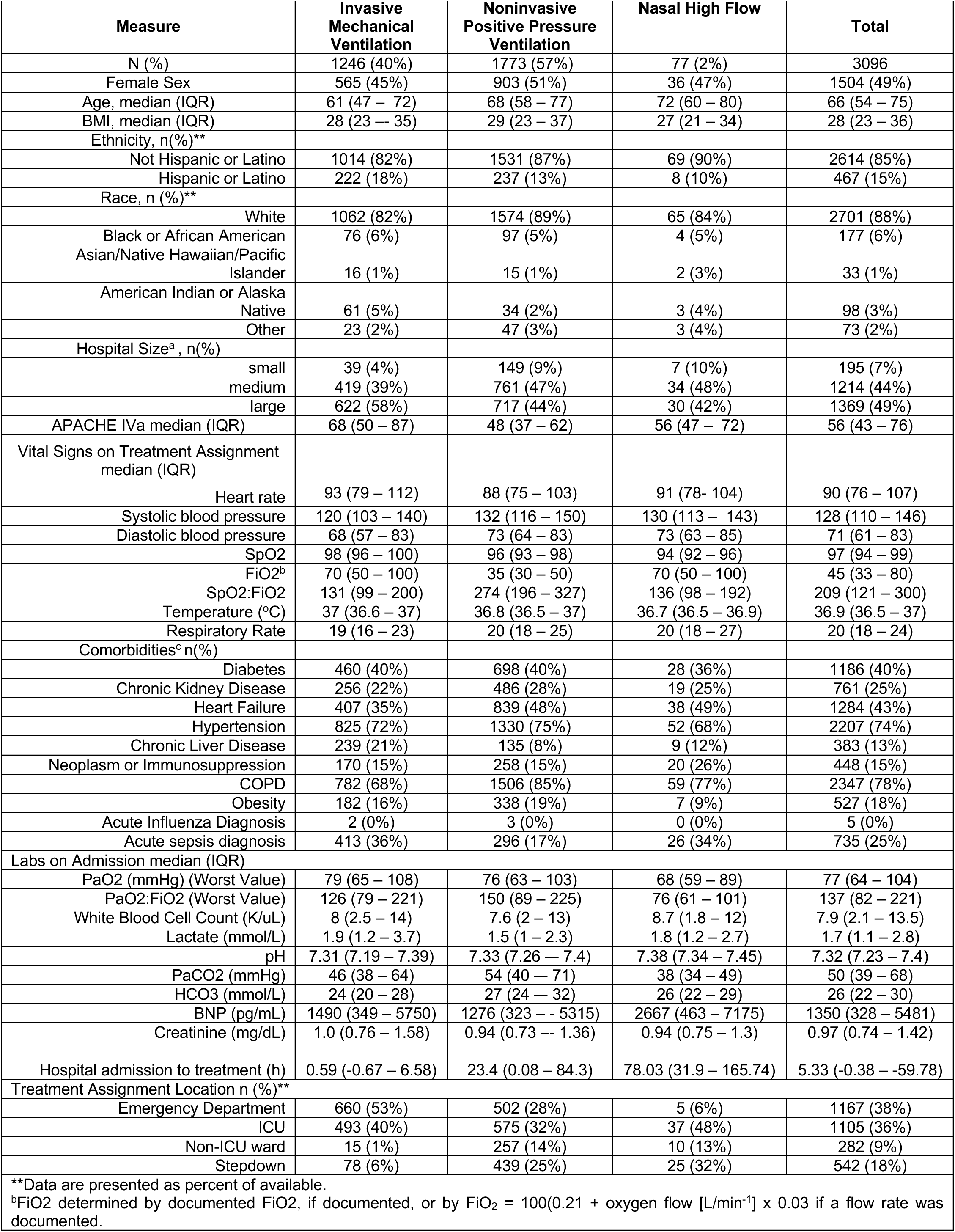
Demographics

## Noninvasive Respiratory Support vs Invasive Mechanical Ventilation Analyses

For model-estimated cumulative incidence curves where patients without clear sequence of support have been assigned their temporally first treatment, discrete covariates other than heart failure, chronic obstructive pulmonary disorder, influenza, and sepsis have been set to the following values: male, not Hispanic or Latino, white, one of the large hospitals (hospital A), hospital admission to the emergency department between January 1, 2018 and June 30, 2018, no vasopressor infusion before treatment, no diabetes, no chronic kidney disease, yes hypertension, no neoplasm/immunosuppression, no chronic liver disease, no obesity, and continuous covariates have been set to their median values (age = 66 years, SpO_2_/FiO_2_ = 200, respiratory rate = 20 breaths/min, BMI = 28.44, transformed hours from hospital admission to first treatment = 1.77). For model-estimated cumulative incidence curves where patients without clear sequence of support have been excluded, the only difference in covariate values is that the transformed hours from hospital admission to first treatment = 1.70. For both types of models, the factors heart failure, chronic obstructive pulmonary disorder, influenza, and sepsis have been allowed to vary.

**Figure E1:**
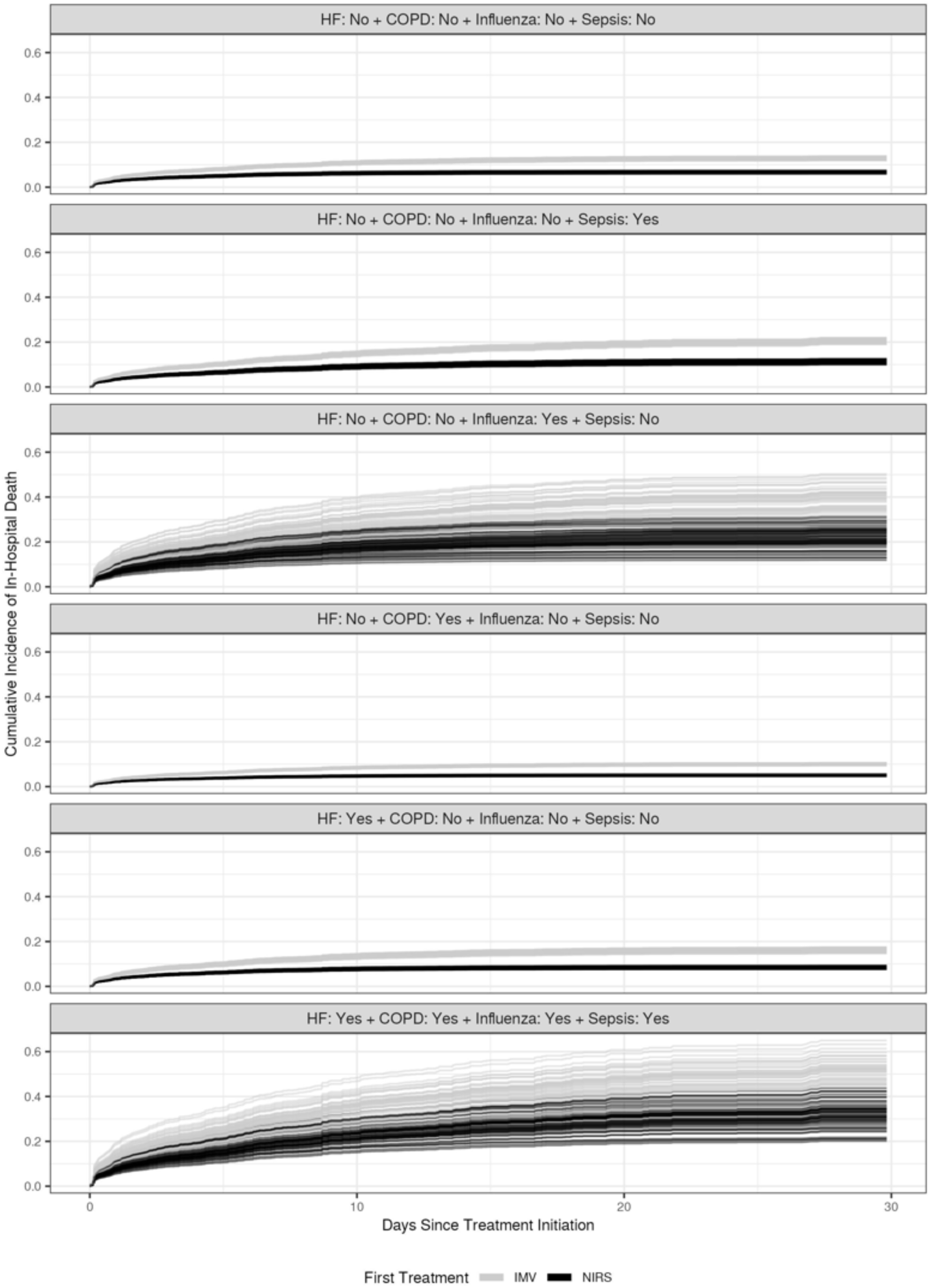
Representative In-Hospital Death Model-Estimated Cumulative Incidence Curves where Patients without Clear Sequence of Support have been Assigned their Temporally First Treatment

**Figure E2:**
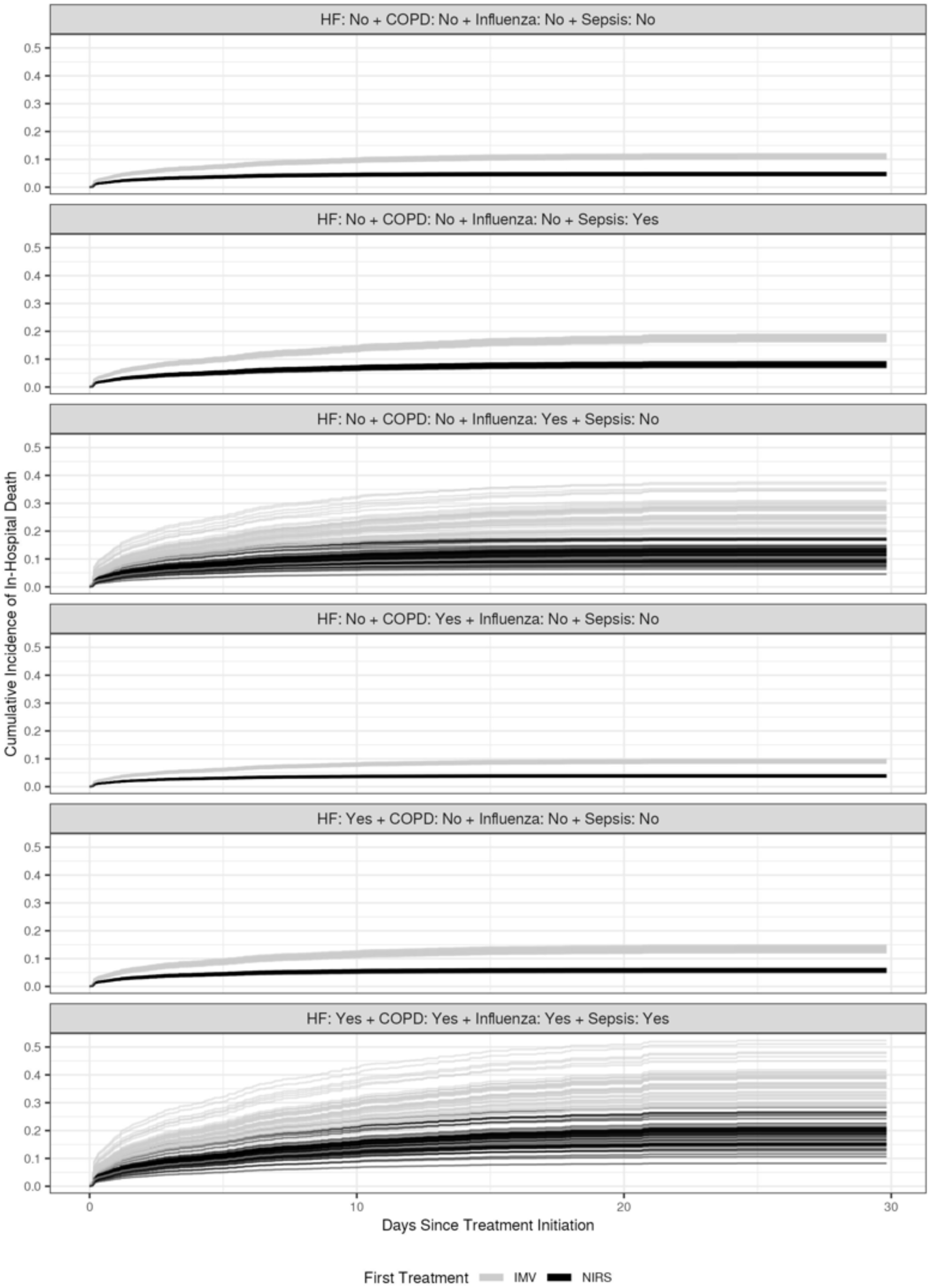
Representative In-Hospital Death Model-Estimated Cumulative Incidence Curves Excluding Patients without Clear Sequence of Support

**Figure E3:**
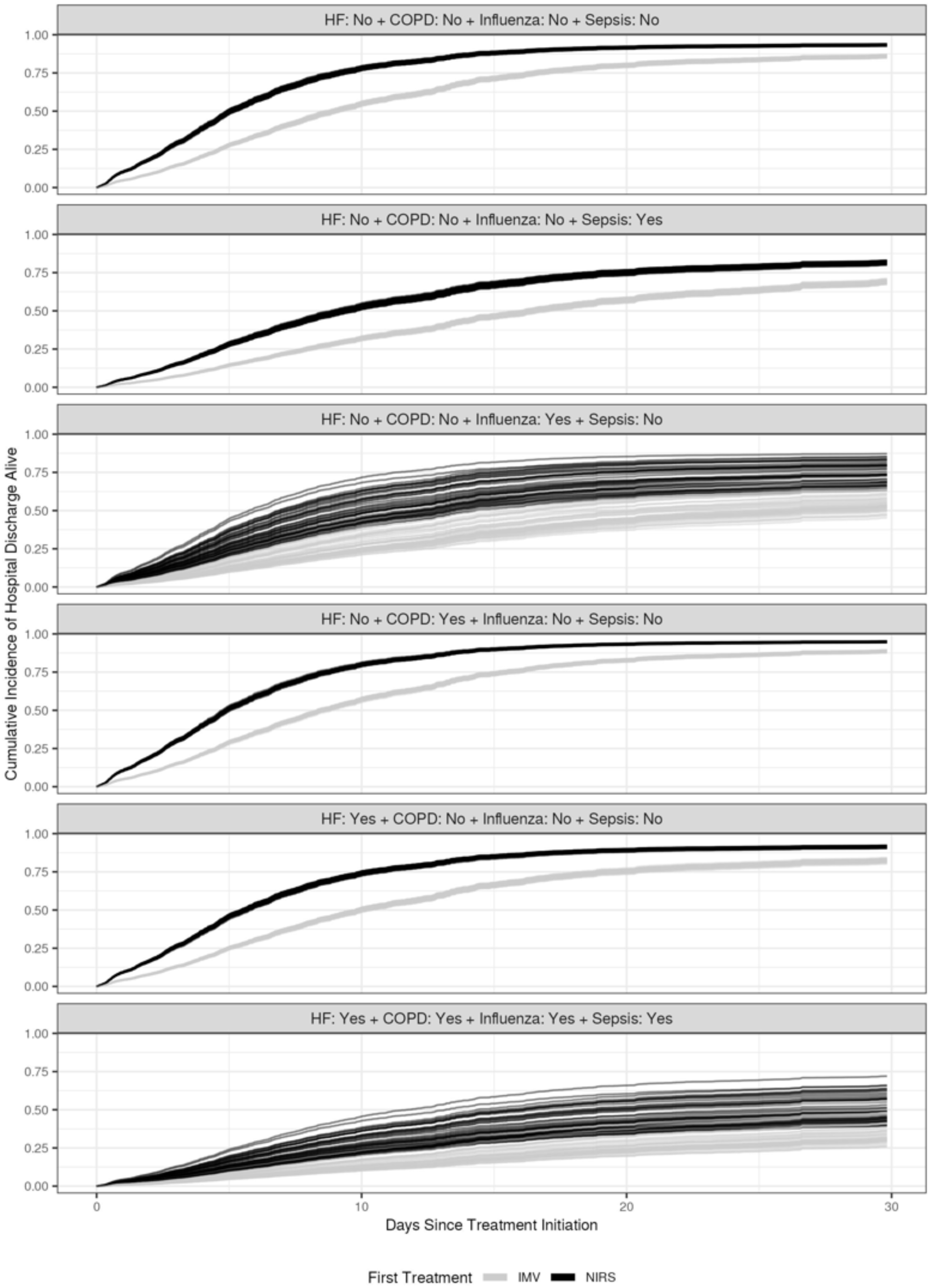
Representative Hospital Discharge Alive Model-Estimated Cumulative Incidence Curves where Patients without Clear Sequence of Support have been Assigned their Temporally First Treatment

**Figure E4:**
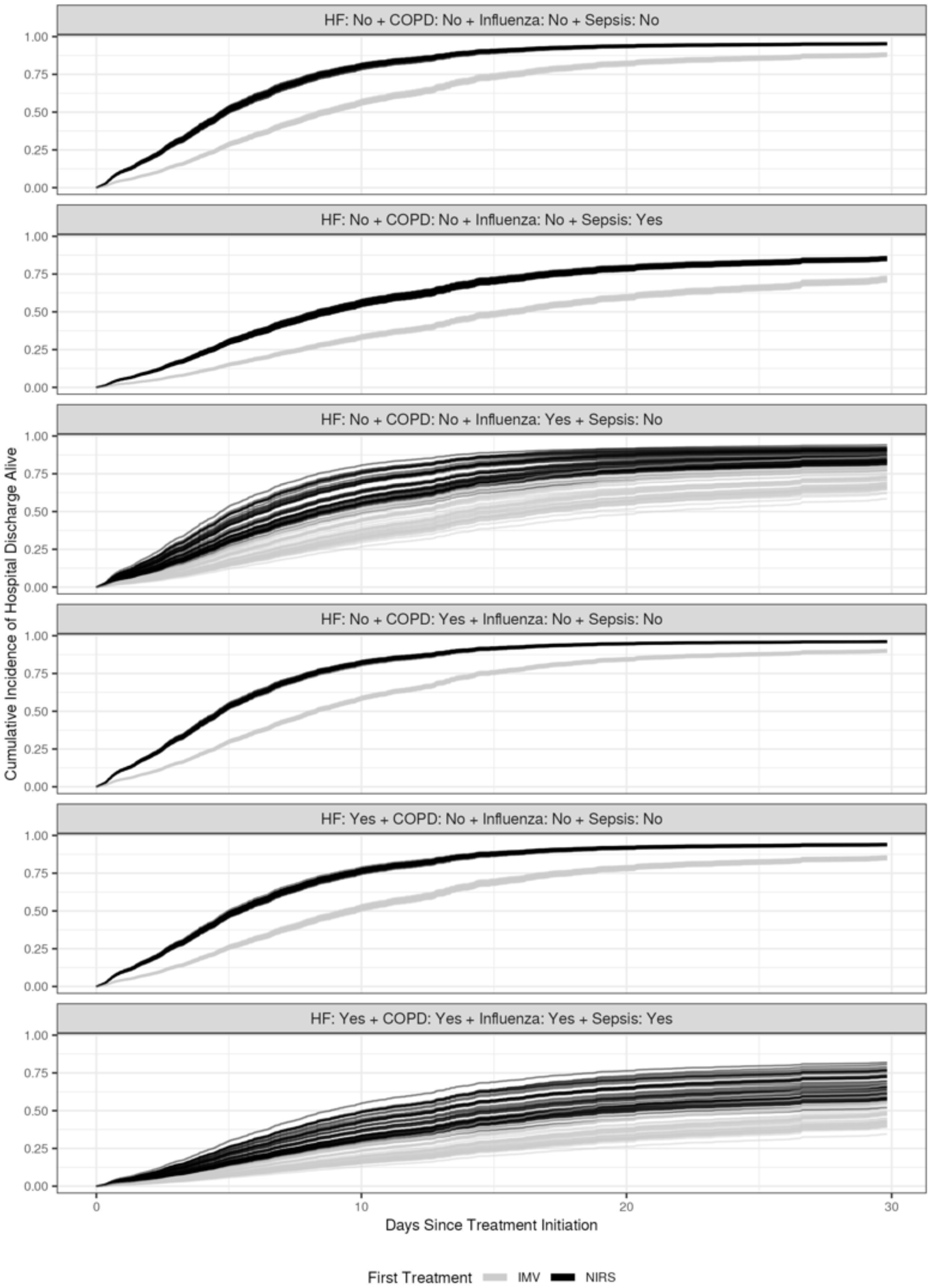
Representative Hospital Discharge Alive Model-Estimated Cumulative Incidence Curves Excluding Patients without Clear Sequence of Support

## Nasal High Flow vs Noninvasive Positive Pressure Ventilation vs Invasive Mechanical Ventilation Analyses

See description of model-estimated cumulative incidence curves for noninvasive respiratory support versus invasive mechanical ventilation.

**Figure E5:**
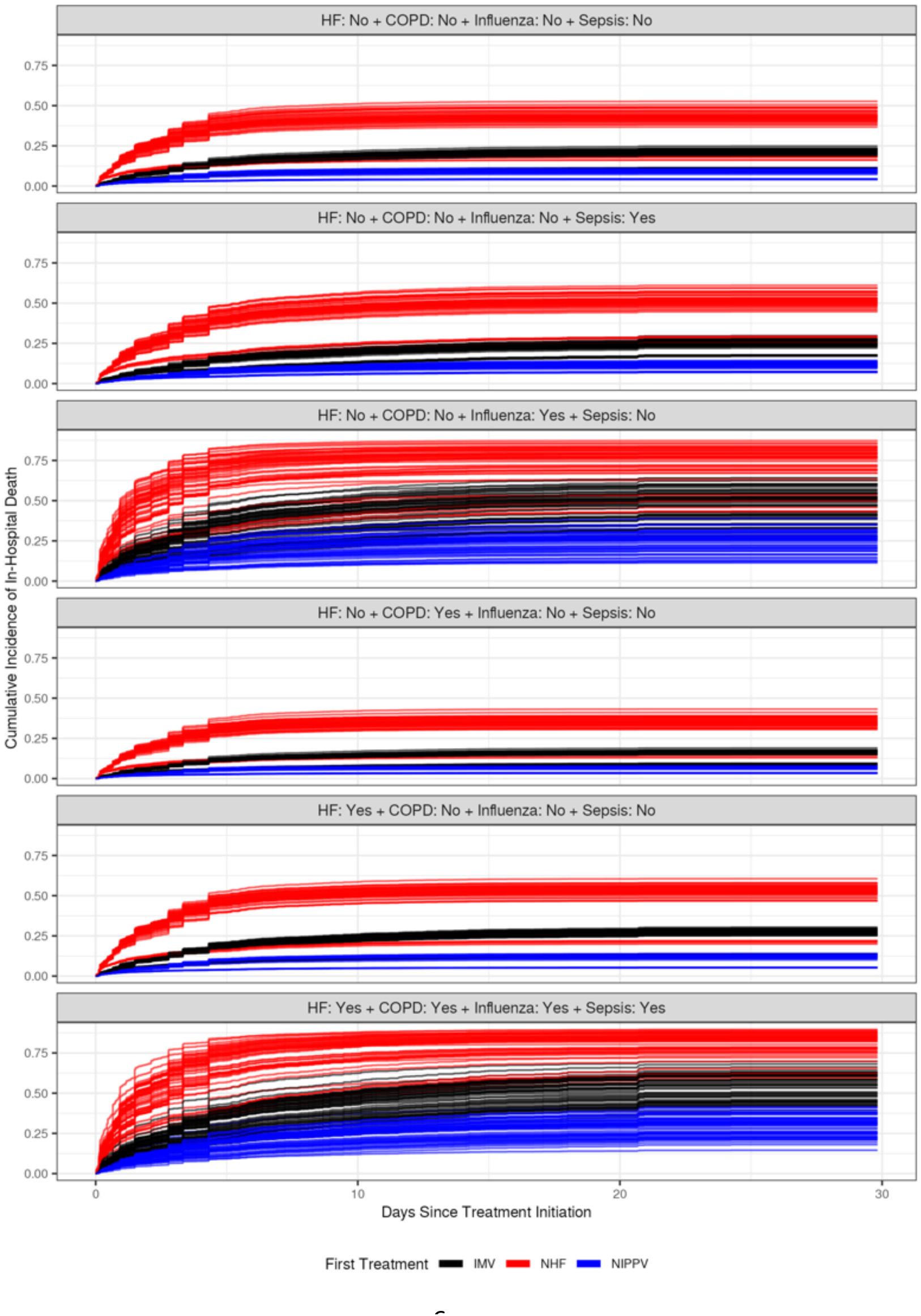
Representative In-Hospital Death Model-Estimated Cumulative Incidence Curves Excluding Patients without Clear Sequence of Support

**Figure E6:**
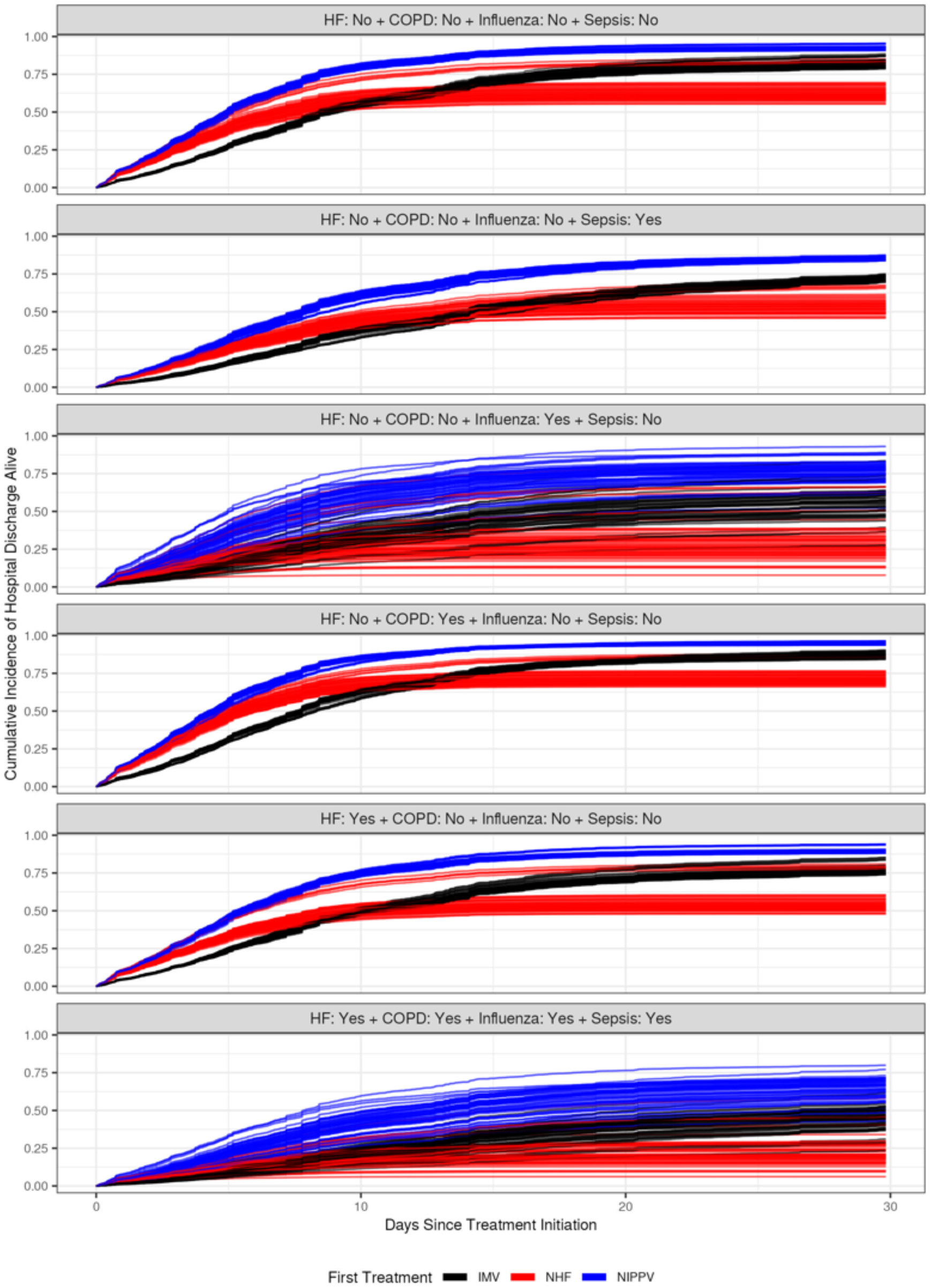
Representative Hospital Discharge Alive Model-Estimated Cumulative Incidence Curves Excluding Patients without Clear Sequence of Support

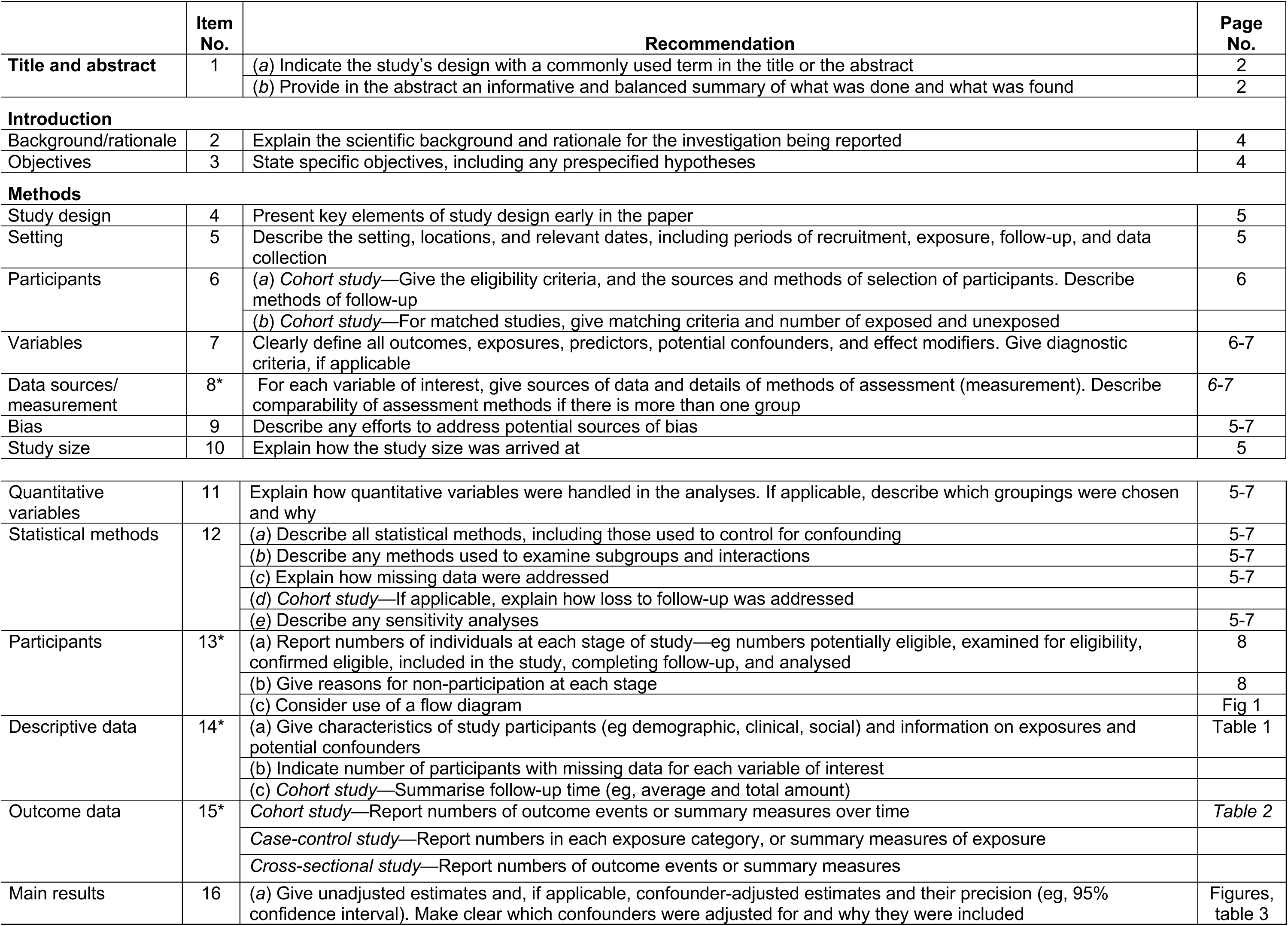
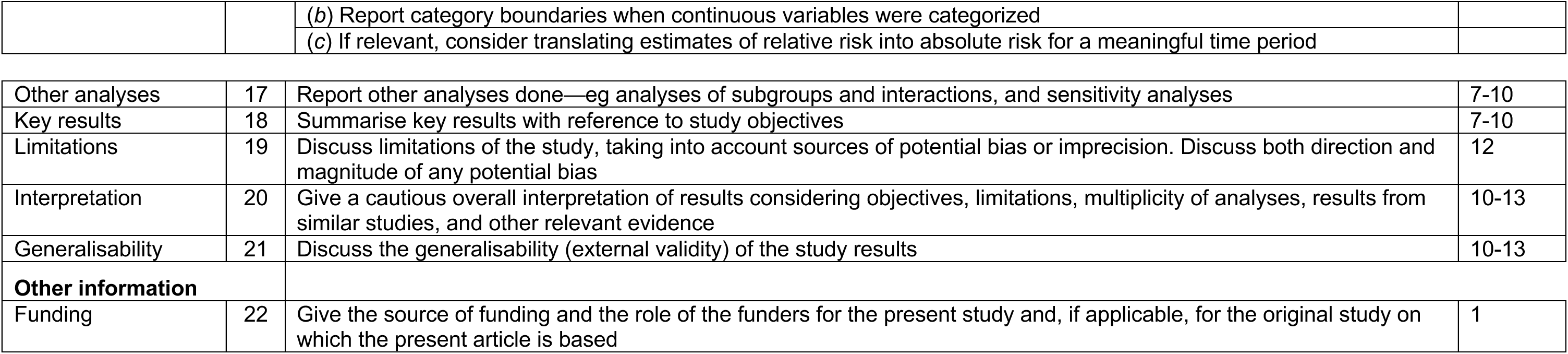
STROBE Checklist

## References

1. Rochwerg B, Brochard L, Elliott MW, Hess D, Hill NS, Nava S, et al. Official ERS/ATS clinical practice guidelines: noninvasive ventilation for acute respiratory failure. The European respiratory journal : official journal of the European Society for Clinical Respiratory Physiology. 2017;50(2).

2. Ferreyro BL, Angriman F, Munshi L, Del Sorbo L, Ferguson ND, Rochwerg B, et al. Association of Noninvasive Oxygenation Strategies With All-Cause Mortality in Adults With Acute Hypoxemic Respiratory Failure: A Systematic Review and Meta-analysis. JAMA : the journal of the American Medical Association. 2020;324(1):57–67.

3. Rochwerg B, Einav S, Chaudhuri D, Mancebo J, Mauri T, Helviz Y, et al. The role for high flow nasal cannula as a respiratory support strategy in adults: a clinical practice guideline. Intensive Care Med. 2020;46(12):2226–37.

4. Pitre T, Zeraatkar D, Kachkovski GV, Leung G, Shligold E, Dowhanik S, et al. Noninvasive Oxygenation Strategies in Adult Patients With Acute Hypoxemic Respiratory Failure: A Systematic Review and Network Meta-Analysis. Chest. 2023.

5. Chaudhuri D, Trivedi V, Lewis K, Rochwerg B. High-Flow Nasal Cannula Compared With Noninvasive Positive Pressure Ventilation in Acute Hypoxic Respiratory Failure: A Systematic Review and Meta-Analysis. Crit Care Explor. 2023;5(4):e0892.

6. Ni YN, Luo J, Yu H, Liu D, Ni Z, Cheng J, et al. Can High-flow Nasal Cannula Reduce the Rate of Endotracheal Intubation in Adult Patients With Acute Respiratory Failure Compared With Conventional Oxygen Therapy and Noninvasive Positive Pressure Ventilation?: A Systematic Review and Meta-analysis. Chest. 2017;151(4):764–75.

7. Shen Y, Zhang W. High-flow nasal cannula versus noninvasive positive pressure ventilation in acute respiratory failure: interaction between PaO2/FiO2 and tidal volume. Crit Care. 2017;21(1):285.

8. Zhao H, Wang H, Sun F, Lyu S, An Y. High-flow nasal cannula oxygen therapy is superior to conventional oxygen therapy but not to noninvasive mechanical ventilation on intubation rate: a systematic review and meta-analysis. Crit Care. 2017;21(1):184.

9. Antonelli M, Conti G, Moro ML, Esquinas A, Gonzalez-Diaz G, Confalonieri M, et al. Predictors of failure of noninvasive positive pressure ventilation in patients with acute hypoxemic respiratory failure: a multi-center study. Intensive Care Med. 2001;27(11):1718–28.

10. Carrillo A, Gonzalez-Diaz G, Ferrer M, Martinez-Quintana ME, Lopez-Martinez A, Llamas N, et al. Non-invasive ventilation in community-acquired pneumonia and severe acute respiratory failure. Intensive Care Med. 2012;38(3):458–66.

11. Demoule A, Girou E, Richard JC, Taille S, Brochard L. Benefits and risks of success or failure of noninvasive ventilation. Intensive Care Med. 2006;32(11):1756–65.

12. Chandra D, Stamm JA, Taylor B, Ramos RM, Satterwhite L, Krishnan JA, et al. Outcomes of noninvasive ventilation for acute exacerbations of chronic obstructive pulmonary disease in the United States, 1998-2008. American Journal of Respiratory and Critical Care Medicine. 2012;185(2):152–9.

13. Walkey AJ, Wiener RS. Use of noninvasive ventilation in patients with acute respiratory failure, 2000-2009: a population-based study. Annals of the American Thoracic Society. 2013;10(1):10–7.

14. Bellani G, Laffey JG, Pham T, Fan E, Brochard L, Esteban A, et al. Epidemiology, Patterns of Care, and Mortality for Patients With Acute Respiratory Distress Syndrome in Intensive Care Units in 50 Countries. JAMA : the journal of the American Medical Association. 2016;315(8):788–800.

15. Bellani G, Laffey JG, Pham T, Madotto F, Fan E, Brochard L, et al. Noninvasive Ventilation of Patients with Acute Respiratory Distress Syndrome. Insights from the LUNG SAFE Study. Am J Respir Crit Care Med. 2017;195(1):67–77.

16. Thille AW, Contou D, Fragnoli C, Cordoba-Izquierdo A, Boissier F, Brun-Buisson C. Non-invasive ventilation for acute hypoxemic respiratory failure: intubation rate and risk factors. Crit Care. 2013;17(6):R269.

17. Brochard L, Slutsky A, Pesenti A. Mechanical Ventilation to Minimize Progression of Lung Injury in Acute Respiratory Failure. Am J Respir Crit Care Med. 2017;195(4):438–42.

18. Lederer DJ, Bell SC, Branson RD, Chalmers JD, Marshall R, Maslove DM, et al. Control of Confounding and Reporting of Results in Causal Inference Studies. Guidance for Authors from Editors of Respiratory, Sleep, and Critical Care Journals. Annals of the American Thoracic Society. 2019;16(1):22–8.

19. Essay P, Mosier J, Subbian V. Rule-Based Cohort Definitions for Acute Respiratory Failure: Electronic Phenotyping Algorithm. JMIR Med Inform. 2020;8(4):e18402.

20. Fisher J, Subbian V, Essay P, Pungitore S, Bedrick E, Mosier J. Acute Respiratory Failure from Early Pandemic COVID-19: Noninvasive Respiratory Support vs Mechanical Ventilation. CHEST Critical Care. 2023.

21. Essay P, Fisher JM, Mosier JM, Subbian V. Validation of an Electronic Phenotyping Algorithm for Patients With Acute Respiratory Failure. Crit Care Explor. 2022;4(3):e0645.

22. Yarnell CJ, Johnson A, Dam T, Jonkman A, Liu K, Wunsch H, et al. Do Thresholds for Invasive Ventilation in Hypoxemic Respiratory Failure Exist? A Cohort Study. Am J Respir Crit Care Med. 2023;207(3):271–82.

23. McCaffrey DF, Griffin BA, Almirall D, Slaughter ME, Ramchand R, Burgette LF. A tutorial on propensity score estimation for multiple treatments using generalized boosted models. Statistics in medicine. 2013;32(19):3388–414.

24. Hawkins D, Weisberg S. Combining the Box-Cox Power and Generalized Log Transformations to Accommodate Nonpositive Responses in Linear and Mixed-Effects Linear Models. South African Statist J. 2017;5:317–28.

25. Putter H, Fiocco M, Geskus RB. Tutorial in biostatistics: competing risks and multi-state models. Statistics in medicine. 2007;26(11):2389–430.

26. Bhattacharjee S, Patanwala AE, Lo-Ciganic WH, Malone DC, Lee JK, Knapp SM, et al. Alzheimer’s disease medication and risk of all-cause mortality and all-cause hospitalization: A retrospective cohort study. Alzheimers Dement (N Y). 2019;5:294–302.

27. Eekhout I, van de Wiel MA, Heymans MW. Methods for significance testing of categorical covariates in logistic regression models after multiple imputation: power and applicability analysis. BMC Med Res Methodol. 2017;17(1):129.

28. Haneuse S, Daniels M. A General Framework for Considering Selection Bias in EHR-Based Studies: What Data Are Observed and Why? EGEMS (Wash DC). 2016;4(1):1203.

29. Botsis T, Hartvigsen G, Chen F, Weng C. Secondary Use of EHR: Data Quality Issues and Informatics Opportunities. Summit Transl Bioinform. 2010;2010:1–5.

30. van der Lei J. Use and abuse of computer-stored medical records. Methods Inf Med. 1991;30(2):79–80.

31. van Buuren S, Groothuis-Oudshoorn K. mice: Multivariate Imputation by Chained Equations in R. Journal of Statistical Software. 2011;45(3):1–67.

32. White IR, Royston P, Wood AM. Multiple imputation using chained equations: Issues and guidance for practice. Statistics in medicine. 2011;30(4):377–99.

33. Team RC. R: A language and environment for statistical computing.: R Foundation for Statistical Computing. Vienna, Austria; 2020.

34. Cefalu M, Ridgeway G, McCaffrey D, A. M. twang: Toolkit for Weighting and Analysis of Nonequivalent Groups. R Package version 2.5 2021 [Available from: https://CRAN.R-project.org/package=twang.

35. T T. A Package for Survival Analysis in R. R package version 3.1-12. 2020.

36. Therneau T, Grambsch P. Modeling Survival Data: Extending the Cox Model. 1 ed. New York: Springer; 2000. 350 p.

37. Kassambara A, Kosinski M, Biecek P. survminer: Drawing Survival Curves using ‘ggplot2.’ R package version 0.4.9 2021 [Available from: https://CRAN.R-project.org/package=survminer.

38. Dahl DB, Scott D, Roosen C, Magnusson A, Swinton J. xtable: Export Tables to LaTeX or HTML. R package version 1.8–4. 2019.

39. Wickham H, Averick M, Bryan J, Chang W, al. e. Welcome to the tidyverse. Journal of Open Source Software. 2019;4(43):1686.

40. Rochwerg B, Granton D, Wang DX, Helviz Y, Einav S, Frat JP, et al. High flow nasal cannula compared with conventional oxygen therapy for acute hypoxemic respiratory failure: a systematic review and meta-analysis. Intensive Care Med. 2019;45(5):563–72.

41. Ni YN, Luo J, Yu H, Liu D, Liang BM, Yao R, Liang ZA. Can high-flow nasal cannula reduce the rate of reintubation in adult patients after extubation? A meta-analysis. BMC pulmonary medicine. 2017;17(1):142.

42. Grieco DL, Menga LS, Raggi V, Bongiovanni F, Anzellotti GM, Tanzarella ES, et al. Physiological Comparison of High-Flow Nasal Cannula and Helmet Noninvasive Ventilation in Acute Hypoxemic Respiratory Failure. Am J Respir Crit Care Med. 2020;201(3):303–12.

43. Tonelli R, Fantini R, Tabbi L, Castaniere I, Pisani L, Pellegrino MR, et al. Early Inspiratory Effort Assessment by Esophageal Manometry Predicts Noninvasive Ventilation Outcome in De Novo Respiratory Failure. A Pilot Study. Am J Respir Crit Care Med. 2020;202(4):558–67.

44. Vieira F, Bezerra FS, Coudroy R, Schreiber A, Telias I, Dubo S, et al. High Flow Nasal Cannula compared to Continuous Positive Airway Pressure: a bench and physiological study. Journal of applied physiology. 2022.

